# Social isolation during the COVID-19 pandemic in Spain: a population study

**DOI:** 10.1101/2022.01.22.22269682

**Authors:** Marina Martinez-Garcia, Emilio Sansano-Sansano, Andrea Castillo-Hornero, Ruben Femenia, Kristof Roomp, Nuria Oliver

## Abstract

Since March of 2020, billions of people worldwide have been asked to limit their social contacts in an effort to contain the spread of the SARS-CoV-2 virus. However, little research has been carried out to date on the impact of such social distancing measures on the social isolation levels of the population. In this paper, we study the impact of the pandemic on the social isolation of the Spanish population, by means of 32,359 answers to a citizen survey collected over a period of 7 months. We uncover (1) a significant increase in the prevalence of social isolation in the population, reaching almost 26%; (2) gender and age differences, with the largest prevalence of isolation among middle-aged individuals; (3) a strong relationship between economic impact and social isolation; and (4) differences in social isolation, depending on the number of COVID-19 protection measures and on the perception of coronavirus infection risk by our participants. Our research sheds quantitative light on the sociological impact of the pandemic, and enables us to identify key factors in the interplay between the deployment of non-pharmaceutical interventions to contain the spread of an infectious disease and a population’s levels of social isolation.

---

**S**ocial isolation is increasingly recognized as an important public health issue. It is defined as “the relative absence of social relationships” (1). Developing instruments to assess the levels of social isolation in a population is of paramount importance given that social engagement –i.e., the lack of social isolation– is a key pillar for good health, as recognized by the World Health Organization (2).

The impact of an individual’s social network on their health and well being has been extensively studied in the literature. Numerous studies have found that social isolation, particularly among older adults, is a risk factor associated with premature death, mainly due to cardiovascular or mental health problems (3–6), with an effect similar to that of obesity, a sedentary lifestyle, substance abuse and cigarette smoking (7). This negative association with health is found even when controlling for other variables, such as socioeconomic factors, perceived loneliness or life habits (7, 8). In recent work by Holt-Lunstad and Steptoe (9), the authors discuss three elements of social relationships (namely function, structure and quality) and their impact on an individual’s health. While they claim that “none of these three elements alone will adequately capture the full scope of social influence on health”, they hypothesize that social isolation “may be a critical component because a weak structural foundation may limit the potential of other social connection factors to have downstream effects on health”.

In the literature, different instruments have been developed to quantify the levels of social isolation in individuals, such as the ENRICHD Social Support Instrument, the Berkman-Syme Social Network Index and the Duke Social Support Indices (10, 11). The Lubben Social Network Scale (LSNS) is one of such instruments. The original version of the LSNS scale consists of 10 items and was originally developed by Lubben in 1988 to assess the social isolation in older adults (12). Recently, a six-item version of the LSNS, called the LSNS-6, was developed by Lubben and Gironda (13–15) and has been validated as a reliable instrument to assess the levels of social isolation both in older (13, 16–18) and younger adults (19).

A related concept to social isolation is that of social capital, which refers to the institutions, relationships and norms that shape the quality and quantity of a society’s social interactions (20). Social capital has both an individual and an aggregate component (21–23). Generally, social capital consists of three dimensions: structural, relational and cognitive. The LSNS-6 measures the relational and structural elements of the social network of an individual by assessing both the social support and the social network aspects, which are part of a person’s individual social capital (24, 25).

Older adults have been reported to be the most vulnerable demographic group to social isolation, mainly due to their reduced physical mobility and general decline in health status, life transitions associated with age (e.g. retirement, loss of a partner, relatives and friends) and ageism in society (11, 26– 28). Beyond the elder population, there are few studies that have assessed the prevalence of social isolation in the general population prior to the COVID-19 pandemic(29, 30).

However, the pandemic has brought social isolation to the public eye not only in the context of older adults, but for all age groups, because of the impact that confinement and social distancing measures have on everyone’s social support structures (31): since March of 2020, billions of people world- wide have been asked to limit their social contacts by physical distancing, home confinements and the temporary closure of many social activities –such as restaurants, bars, gyms, workplaces, soccer stadiums, museums, cinemas and theaters– in an effort to contain the spread of the SARS-CoV-2 virus.

Despite the importance of social isolation in this context, there are few population studies published to date that have analyzed the impact of the COVID-19 pandemic on a population’s levels of social isolation (32–34). One of the most relevant pieces of previous work is by O^*/*^Sullivan et al. (35) where they studied the impact of the pandemic on the social isolation of the general population of 101 different countries, mainly from the United States (40%) and the UK (21%). The authors collected the answers to the LSNS-6 survey from 14,302 participants aged over 18 years old (M=53 years, SD=17.6) before and during the coronavirus pandemic. They found a 13% increase in the prevalence of social isolation during the pandemic. The most important factors associated with social isolation –both before and during the coronavirus pandemic– are a lack of financial resources to meet the participants’ needs, their self-rated physical and mental health, living alone not by choice, poor sleep, low physical activity and alcohol consumption. The most cross-cutting aspects correlated with social isolation were economic and mental health factors. During the pandemic, rural-town dwellers were found to be significantly at larger risk of social isolation when compared to city dwellers.

Given previous research on this topic, in this paper we focus on studying the levels of social isolation in the Spanish population during the COVID-19 pandemic by means of the LSNS-6 instrument, delivered as part of an online, anonymous survey called the COVID19ImpactSurvey (36) with over 720,000 answers collected since March of 2020. The answers to the survey have been previously analyzed for research purposes and extensively used by public authorities, the media and citizens in Spain to shed light on the impact of the COVID-19 pandemic on people’s behaviors, perceptions and lives (37, 38). While the original survey did not include any question to assess the levels of social isolation of the respondents, on June 6th 2021 the LSNS-6 questions were added to the survey and they have been deployed since. Thus, we analyze a sample of 32,359 answers collected in Spain for the time period between June 6th, 2021 and December 16th, 2021. On the one hand, Spain was one of the most affected countries in the early phases of the COVID-19 pandemic during the Spring of 2020. In an effort to contain the spread of the virus, the government implemented very restrictive population lock-downs that brought the country to a halt for several weeks, with tremendous economic and social implications. On the other hand, Spain is a country with a highly social culture, with low levels of individualism and where families are a fundamental pillar in most of the Spaniard’s lives (39–42).

People in Spain tend to handle their personal problems through their family, relying on relatives for support when facing difficult situations, such as a pandemic. In our analysis, we aim to answer four research questions on the interplay between the economic and psychological impact of the COVID-19 pandemic and the self-reported behavior and levels of social isolation of the Spanish population, namely:

**RQ1**: What is the distribution and the temporal evolution of the levels of social isolation in the Spanish population during six months of the COVID-19 pandemic?

**RQ2**: What is the relationship between social isolation and the economic impact of the pandemic on individuals?

**RQ3**: What is the relationship between social isolation and the psychological impact of the pandemic on individuals?

**RQ4**: Are there differences in the levels of social isolation between those with low perceptions of infection risk associated with common daily activities and those who do not adopt any COVID-19 protective measures when compared to the rest of the sample?

The paper is structured as follows: in the following section, we present the Results of analyzing the survey answers to address the four previously stated research questions. Next, we Discuss the results and draw key conclusions from our research. Finally, we describe our Materials and Methods.

## Results

In this section, we describe the results of our analyses to answer the previously formulated research questions, RQ1 to RQ4.

We analyze 32,359 answers to the COVID19ImpactSurvey (36), which two of the authors designed and launched on March 28th 2020. Our results are based on analyzing a subset of 17 questions of this survey as per Table 3 in the Supporting Information (S.I.). We focus on the answers collected between June 6th, 2021 and the 16th of December, 2021 in Spain, namely 32,359 answers. Respondents were required to be at least 18 years old.

The gender and age distributions of the collected sample are not proportional to those of the general population of Spain, as shown in Table 5. Thus, we follow the methodology described in Oliver et al. (36) and re-weigh the answers, such that the resulting gender and age distributions match the official statistics in Spain in 2020. We also filter entries with inconsistent answers (5.19% of answers), entries that appear to have been answered too fast or too slow (3.34% of answers), and entries not providing age or male/female sex information (0,81%). In total, we discard 9.34% of the answers. We refer to the Materials and Methods section for additional details on the survey data processing. All answers are categorical or binary, thus we report the percentage of participants who selected each response and compute the 95% confidence intervals (CI) through the margin of error. All tests of significance have been done by means of t-test or ANOVA as appropriate and post-hoc comparisons have been performed using Tukey tests.

### RQ1: Distribution and temporal evolution of social isolation

#### Sample Characteristics

Complete LSNS-6 scores were available for 27,898 participants 51.49% were women and 48.51% were men in the re-weighted sample. The mean age of the study sample was 42.45 years. The mean LSNS-6 score of the study population was *μ*_*LSNS*−6_ = 15.23 (SD = 0.043). The average LSNS-6 scores were 7.66 (SD=0.041) and 17.89 (SD =0.036) for socially isolated (*μ*_*LSNS*−6_<12) and socially integrated (*μ*_*LSNS*−6_ ≥ 12) individuals, respectively.

#### Prevalence of Social Isolation

The prevalence of social isolation was *prev*_*iso*_ = 25.98% (SD=0.003) across all ages in the sample. Social isolation was more prevalent in men when compared to women (men: *prev*_*iso*_ = 26.53%, women: *prev*_*iso*_ = 25.47%, yet we did not identify a statistically significant difference in the prevalence between genders via a *χ*^2^ test. Conversely, the LSNS-6 average scores among men were larger than among women (men: *μ*_*LSNS*−6_ = 15.29, SD=0.074; women: *μ*_*LSNS*−6_ = 15.17, SD=0.047).

The demographic group with the largest average LSNS-6 scores is the youth (18 to 29 years old), followed by the elderly (aged 60+ years old) and those aged 30-39 years old. Interestingly, the middle-aged (40 to 59 years old) group exhibits the lowest LSNS-6 values, yielding an “U”-shaped curve of LSNS-6 scores, as shown in Figure 1. In our data data, 29.54% of the individuals aged 50 to 59 years old report being socially isolated, a figure significantly larger than what has been previously presented in the literature prior to the pandemic (30).

**Fig. 1.**
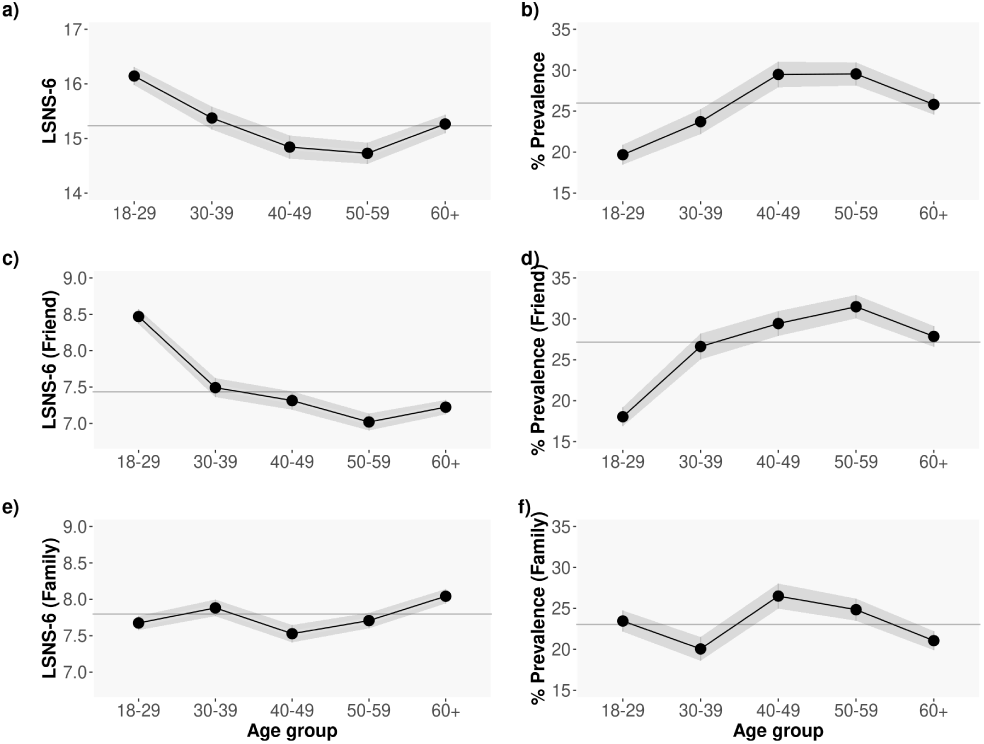
Average **a)** LSNS-6, **c)** LSNS-6 Friends and **e)** LSNS-6 Family scores per age group; Proportion of non isolated individuals per age group, defining isolation as an **b)** LSNS-6 score < 12, **d)** LSNS-6 Friend score < 6, and **f)** SNS-6-Family score < 6. The greyed area shows the 95% CI. The grey continuous line correspond to the overall average LSNS-6 score for all the age groups: *μ*_*LSNS-*6_ = 15.23, *SD* = 0.043; *prev*_*iso*_ = 25.98%, *SD* = 0.003; *μ*_*LSNS-*6 *fri*_ = 7.43, *SD* = 0.026; *μ*_*LSNS-*6 *fam*_ = 7.80, *SD* = 0.025; *prev*_*iso fri*_ = 27.16%; *prev*_*iso fam*_ = 23.04%.

To shed light on the contributions of friends vs family relationships in the overall LSNS-6 score, Figures 1 c), d), e) and f) show the average LSNS-6 Friend and Family scores and average prevalence of social isolation by age group. As can be seen in the Figure, the LSNS-6 scores in the youth (18-29 years old) are strongly reliant on the Friends component whereas in the elderly (60+ years old) they are more dependent on the Family component: *μ*_*LSNS*−6 *fam*_ : 7.67 in the youth vs 8.04 in the elderly; *μ*_*LSNS*−6 *fri*_ : 8.47 in the youth vs 7.22 in the elderly.

The temporal evolution of the average LSNS-6 scores across time is shown in Figure 2 a). As depicted in the Figure, the average values are somewhat stable over time except for a notable decrease in the LSNS-6 scores collected during week 27 (5th to 11th of July, 2021) and an increase in the LSNS-6 scores corresponding to week 38 (20th to 26th of September, 2021), which might be due to the return to work and school after the summer holidays. A LOESS curve fit reveals a growing trend of the average LSNS-6 scores starting in week 43, which might be reflective of the return to the “new normality” and the *pandemic fatigue* in the population (37).

**Fig. 2.**
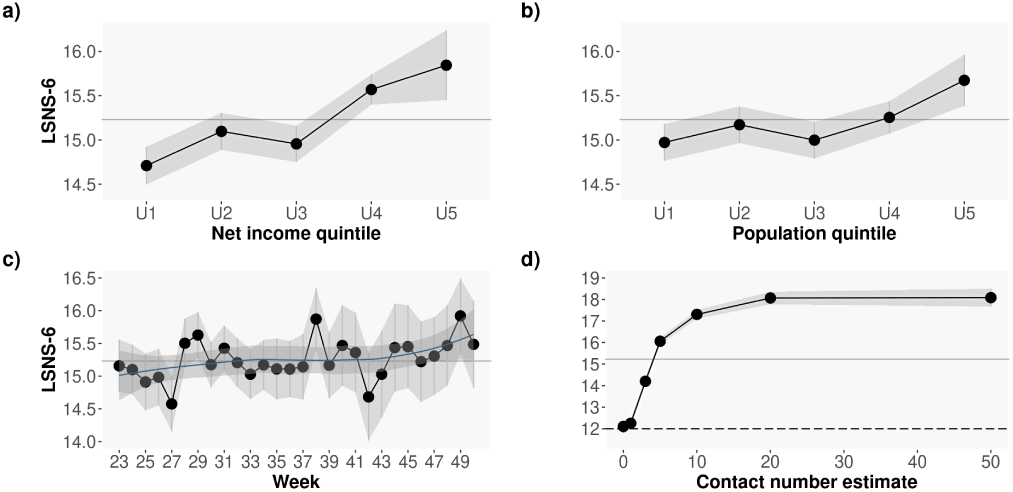
Average LSNS-6 score by **a)** income and **b)** population (in thousands), both obtained from the 2020 Spanish official statistics (INE) data according to the participants’ reported postal code; **c)** Average LSNS-6 score by week of the year: from June 6th to December 16th, 2021; **d)** Average LSNS-6 score for the *Number of close contacts in the last 7 days (Q9)* responses. The greyed area shows the 95% CI. The grey continuous line correspond to the overall average LSNS-6 for the entire sample: *μ*_*LSNS-*6_ = 15.23, *SD* = 0.043.

#### Associations between social isolation and socio-demographic and economic factors

Previous work has identified significant correlations between social isolation, income and population density. In our data, we observe a similar pattern to that reported in the literature: Figures 2 b) and c) depict the average LSNS-6 scores for all the participants living in postal codes grouped by their associated income (b) and population (c) as reported by the Spanish National Institute of Statistics (data for 2020). As seen in the Figure, the larger the levels of isolation (i.e. the lower the LSNS-6 score), the lower the income and the lower the population: those living in poorer areas and those living in smaller municipalities exhibit significantly lower LSNS-6 scores than those living in more affluent or/and populated regions.

Logistic regression analysis confirmed associations of age, number of people living in the household, average income and population of the respondents’ zip code with social isolation, as depicted in Table 2. In the Table, the income is divided in three bins: low [12,201-22,064 € /year]; medium [22,065-25,414 € /year]; and high [25,415-60,263 € /year], and so is the population: low [119-44,320 inhabitants]; medium [44,321-312,004 inhabitants]; and high [312,005 -5,226,965 inhabitants].

**Table 1.** **Logistic regression analysis of social isolation with age, gender, number of people in the home, income and population as independent variables. The asterisks denote statistical significance of the coefficient in the logistic model,** 0.001 <^*\*\*\**^, 0.01 <^*\*\**^, 0.05 <^*\**^

|  | Odds ratio | Int low | Int sup |
| --- | --- | --- | --- |
| Age Group: Basal value 18-29 |  |  |  |
| 30-39 ** | 1.25 | 1.09 | 1.43 |
| 40-49*** | 1.72 | 1.52 | 1.95 |
| 50-59*** | 1.70 | 1.51 | 1.92 |
| 60+*** | 1.36 | 1.21 | 1.54 |
| Gender: Basal value Female |  |  |  |
| Male | 1.03 | 0.96 | 1.11 |
| Number people household: Basal value 1 |  |  |  |
| 2*** | 0.70 | 0.62 | 0.79 |
| 3*** | 0.73 | 0.65 | 0.83 |
| 4+*** | 0.65 | 0.57 | 0.74 |
| Income: Basal value Low |  |  |  |
| Medium | 1.00 | 0.92 | 1.10 |
| High*** | 0.68 | 0.67 | 0.84 |
| Population: Basal value Low |  |  |  |
| Medium** | 0.87 | 0.80 | 0.96 |
| High | 1.02 | 0.92 | 1.14 |

**Table 2.** **Logistic regression analysis of social isolation with age, gender, number of people in the home, income, population, economic and psychological impact as independent variables. The asterisks denote statistical significance of the coefficient in the logistic model, p-values** 0.001 <^***^, 0.01 <^**^, 0.05 <^*^

|  | Odds ratio | Int low | Int sup |
| --- | --- | --- | --- |
| Age Group: Basal value 18-29 |  |  |  |
| 30-39*** | 1.29 | 1.12 | 1.48 |
| 40-49*** | 1.83 | 1.61 | 2.08 |
| 50-59*** | 1.89 | 1.67 | 2.14 |
| 60+*** | 1.59 | 1.39 | 1.81 |
| Gender: Basal value Female |  |  |  |
| Male | 1.07 | 1.00 | 1.16 |
| Number people household: Basal value 1 |  |  |  |
| 2*** | 0.71 | 0.63 | 0.80 |
| 3*** | 0.73 | 0.64 | 0.83 |
| 4+*** | 0.65 | 0.57 | 0.74 |
| Income: Basal value Low |  |  |  |
| Medium | 1.02 | 0.93 | 1.12 |
| High*** | 0.77 | 0.68 | 0.86 |
| Population: Basal value Low |  |  |  |
| Medium** | 0.86 | 0.78 | 0.94 |
| High | 1.00 | 0.90 | 1.11 |
| Economic impact: Basal value None |  |  |  |
| Positive* | 0.89 | 0.79 | 1.00 |
| Mild*** | 1.18 | 1.08 | 1.30 |
| Severe*** | 1.90 | 1.58 | 2.27 |
| Psychological impact: Basal value None |  |  |  |
| Mild** | 0.84 | 0.74 | 0.95 |
| Severe*** | 1.39 | 1.27 | 1.51 |

**Table 3.** COVID19ImpactSurvey questions analysed in our study.

| Question | Possible answers |
| --- | --- |
| **Demographic and Household information** |  |
| Q1. What is your age range? | [18-20; 21-29; 30-39; 40-49; 50-59; 60-69; 70-79; 80+] |
| Q2. What is your gender? | [Male; female; another gender] |
| Q3. Postal code/Zip code | Text entry |
| Q4. Type of home | [Single family house; apartment/flat; shared apartment/flat; other shared accommodation; other] |
| Q5. Number of people in the home (including you) | [1; 2; 3; 4; 5+] |
| Q6. Age(s) of people in the home (excluding you, check all that apply) | [10 or less; 11-20; 21-29; 30-39; 40-49; 50-59; 60-69; 70-79; 80+] |
| **Social Behaviour** |  |
| Q9. In the last seven days, approximately how many different people that live outside your home have you had close contact with? (meaning more than 15 minutes and a distance closer than 2 meters) | [No one; 1-2; 3-4; 5-9; 10-19; 20-49; 50+] |
| **LSNS-6** |  |
| Q11. How many relatives do you see or hear from at least once a month? (considering the people to whom you are related by birth, marriage, adoption, etc.) | [No one; 1; 2; 3-4; 5-8; 9+] |
| Q12. How many relatives do you feel at ease with that you can talk about private matters? | [No one; 1; 2; 3-4; 5-8; 9+] |
| Q13. How many relatives do you feel close to such that you could call on them for help? | [No one; 1;2;3-4;5-8;9+] |
| Q14. How many of your friends do you see or hear from at least once a month? (considering all of your friends including those who live in your neighborhood) | [No one; 1;2;3-4;5-8;9+] |
| Q15. How many friends do you feel at ease with that you can talk about private matters? | [No one; 1;2;3-4;5-8;9+] |
| Q16. How many friends do you feel close to such that you could call on them for help? | [No one; 1;2;3-4;5-8;9+] |
| Q19. Do you believe that the measures the government has taken are enough to contain the spread of coronavirus? | [No, but should be stricter; Yes, are about right; Yes, but are too strict; Prefer not to respond; I do not know] |
| **Economic Impact** |  |
| Q20. What kind of economic impact has the coronavirus had on you? (check all that apply) | [I lost my job; I lost my savings; I cannot pay my rent or mortgage anymore; I cannot afford to buy food; I have lost most or all of my income; My business is in danger of bankruptcy; My employer is in danger of bankruptcy; My employer has reduced by working hours due to lack of demand; I have a new job or business opportunity; I have significantly increased my savings or reduced my debt because I am spending less; None of the above] |
| Q22. In the last month, have you received any of the following economic assistance from your government? (check all that apply) | [Retirement (pension or social security); Unemployment benefit (temporary); Unemployment benefit (I lost my job); Universal Basic Income; Using the moratorium to reduce my taxes, mortgage, rent or other debt payments; One-time bonus to cover COVID expenses or stimulate the economy; Social housing or subsidised rent; Assistance for my business that I have to pay back (like a loan); Assistance for my business that I do not have to pay back; Disability benefits; Other type of welfare (due to poverty, illness or other reason); None of the above] |
| **Perception of infection risk** |  |
| Q21. Which of the following activities do you think can be done with a low risk of coronavirus infection? (check all that apply) | [Practising individual sports; Having friends visit you at home; Attending religious services with limited seating; Attending school like in some European countries; Going to small businesses with appointment (hairdresser, etc); Going to small shops while maintaining a safe distance; Having drinks at a bar on an open terrace with a group of people; Going to restaurants with limited seating; Receiving treatment at a hospital; Taking public transportation with space between seating; Going to the beach; Travelling by air; None of the above] |
| **COVID-19 infection protection measures** |  |
| Q26. Do you take any of the following measures to prevent the transmission of the coronavirus? (check all that apply) | [I wear a mask as much as possible; I avoid crowded situations; I don't shake hands, give hugs or kisses to anyone who live outside my home; I regularly disinfect/wash my hands; I keep my physical distance of at least 1.5 meters (6 feet) from others; I limit the number of people that I am in close contact with When indoors; I make sure there is good ventilation; I have installed my government's contact tracing app on my phone; I would be willing to get vaccinated immediately when the coronavirus vaccine is available; None of the above] |
| **Psychological Impact** |  |
| Q31. Have you noticed a significant increase in your home in any of the following areas that you consider damaging? (check all that apply) | [High level of anxiety; High level of stress; High level of loneliness; High level of sadness; Loud arguments or fights with other members of the home; Excessive consumption of alcohol; Excessive consumption of drugs (prescription or other); Excessive use of technology by adults (tablet, phone, TV); Excessive use of technology by children (tablet, phone, TV); I have not noticed a harmful increase in these areas; I prefer not to answer] |

### Relationship between the contact number estimate and LSNS-6 scores

Question 9 in the survey (see Table 3 in S.I.) asks participants to estimate the number of different people from outside the home they have had a close contact with in the past week. This question is meant to assess the intensity of social relationships in the survey respondents. Figure 2 d) shows the relationship between the reported number of close contacts and the LSNS-6 scores of the participants in our sample. As seen in the Figure, the larger the number of close contacts, the higher the LSNS-6 scores, i.e. the lower the probability of social isolation.

### RQ2: Social isolation and economic impact of the pandemic

We address RQ2 by analyzing the answers to questions Q20 and Q22 in the survey (see Tables 3 and 4 in the S.I.), as described next.

**Table 4.** Independent and dependent variables used in our study.

| Variable/Values | Explanation |
| --- | --- |
| *Socially Isolated* |  |
| 1 | LSNS-6 score < 12 |
| 0 | LSNS-6 score ≥ 12 |
| *Age (Q1)* |  |
| 18-29 | 18 to 29 years old |
| 30-39 | 30 to 39 years old |
| 40-49 | 40 to 49 years old |
| 50-59 | 50 to 59 years old |
| 60+ | 60 or more years old |
| *Gender (Q2)* |  |
| Male | Male respondents |
| Female | Female respondents |
| *Number of people in the home (Q5)* |  |
| "1" | People who answered "1" |
| "2" | People who answered "2" |
| "3" | People who answered "3" |
| "4+" | People who answered "4+" |
| *Close contact in the last 7 days (Q9)* |  |
| No one | People who answered "No one" |
| 1-2 | People who answered "1-2" |
| 3-4 | People who answered "3-4" |
| 5-9 | People who answered "5-9" |
| 10-19 | People who answered "10-19" |
| 20-49 | People who answered "20-49" |
| 50+ | People who answered "50+" |
| *Economic Impact (Q20)* |  |
| None | No economic impact is reported |
| Mild | I lost my job; I lost part or all of my savings; I have lost most or all of my income<br>My business is in danger of bankruptcy ; My employer is at risk of bankruptcy ; My employer has reduced my hours due to lack of demand |
| Severe | I can't afford to buy food; I can't pay my rent or mortgage anymore |
| Positive | I have a new job or business opportunity; I have significantly increased my savings |
| *Government eco. assistance (Q22)* |  |
| None | People who do not receive any government support |
| Any | People who receive at least one of the following: Assistance for my business that I have to pay back (like a loan); Assistance for my business that I do not have to pay back; Disability benefits; Assistance cash, Assistance loan, One-time bonus to cover COVID expenses or stimulate the economy; Using the moratorium to reduce my taxes, mortgage, rent or other debt payments; Social housing or subsidised rent; Unemployment benefit (I lost my job); Unemployment benefit (temporary); Universal Basic Income |
| *Activities with perceived low risk of COVID-19 infection (Q21)* |  |
| Classification of activities |  |
| **Essential:** | Receiving treatment at a hospital; Attending school like in some European countries; Going to small shops while maintaining a safe distance; Taking public transportation with space between seating; Going to small businesses with appointment (hairdresser, etc); |
| **Non-essential:** | Going to restaurants with limited seating; Going to the beach; Attending religious services with limited seating; Having drinks at a bar on an open terrace with a group of people; Practising individual sports; Travelling by air; Having friends visit you at home; |
| Grouping |  |
| None/Only essential | No activity, or only those that are essential, can be safely reopened: People who answered Yes to open at least one essential and "No" to all other activities |
| Other activities | People who answered Yes to open at least one essential activity, and answered Yes to at least one of the other activities. |
| *COVID-19 Protection Measures (Q26)* |  |
| None | People who are not taking any health safety measures *None of the above* |
| Some | People who answered "Yes" to at most 4 health safety measures |
| Several | People who answered "Yes" to 5 or more safety measures |
| *Psychological Impact (Q31)* |  |
| None | No psychological impact is reported |
| Mild | Excessive use of technology by adults (tablet, phone, TV); Excessive use of technology by children (tablet, phone, TV) |
| Severe | High levels of anxiety; High levels of stress; High levels of loneliness; High levels of sadness; Loud arguments or fights with other members of the home; Excessive consumption of alcohol; Excessive consumption of drugs (prescription or other) |

**Table 5.** Distribution of answers per social isolation values.

| Variable | Responses |  | LSNS-6 |  | Prevalence |  |
| --- | --- | --- | --- | --- | --- | --- |
|  | raw | reweigh | raw | reweigh | raw | reweigh |
| *Socially Isolated* |  |  |  |  |  |  |
| 1 | 25.54 % | 25.98% | 7.81 | 7.66 | - | - |
| 0 | 74.46 % | 74.02% | 17.73 | 17.89 | - | - |
| *Age (Q1)* |  |  |  |  |  |  |
| 18-29 | 21.58% | 15.03% | 15.95 | 16.14 | 19.90% | 19.68% |
| 30-39 | 17.17% | 15.38% | 15.25 | 15.37 | 24.03% | 23.72% |
| 40-49 | 20.37% | 20.05% | 14.76 | 14.84 | 29.14% | 29.48% |
| 50-59 | 19.97% | 18.15% | 14.62 | 14.73 | 29.78% | 29.55% |
| 60+ | 20.91% | 31.40% | 15.36 | 15.27 | 25.03% | 25.82% |
| *Gender (Q2)* |  |  |  |  |  |  |
| Male | 28.22% | 48.51% | 15.36 | 15.29 | 25.94% | 26.53% |
| Female | 71.78% | 51.49% | 15.13 | 15.18 | 25.38% | 25.47% |
| *Number of people in the home (Q5)* |  |  |  |  |  |  |
| 1 | 10.69% | 13.13% | 14.59 | 14.38 | 28.54% | 30.77% |
| 2 | 33.18% | 36.53% | 15.19 | 15.29 | 25.11% | 25.25% |
| 3 | 26.57% | 24.60% | 15.04 | 15.09 | 26.01% | 26.22% |
| 4+ | 29.56% | 25.74% | 15.57 | 15.76 | 24.31% | 24.03% |
| *Close contact in the last 7 days (Q9)* |  |  |  |  |  |  |
| No one | 9.50% | 9.24% | 12.31 | 12.10 | 45.02% | 46.00% |
| 1-2 | 16.41% | 15.79% | 12.49 | 12.25 | 42.44% | 43.98% |
| 3-4 | 20.68% | 20.07% | 14.28 | 14.20 | 28.09% | 28.93% |
| 5-9 | 23.19% | 22.73% | 16.07 | 16.06 | 17.63% | 18.52% |
| 10-19 | 16.64% | 17.38% | 17.32 | 17.30 | 14.29% | 15.21% |
| 20-49 | 7.77% | 8.18% | 18.02 | 18.07 | 12.26% | 11.95% |
| 50+ | 5.81% | 6.60% | 17.51 | 18.08 | 18.70% | 17.28% |
| *Economic Impact (Q20)* |  |  |  |  |  |  |
| None | 58.11% | 60.07% | 15.42 | 15.40 | 24.19% | 24.79% |
| Mild | 22.62% | 20.98% | 14.56 | 14.71 | 29.48% | 29.30% |
| Severe | 3.85% | 3.68% | 12.89 | 13.09 | 42.30% | 41.39% |
| Positive | 15.41% | 15.27% | 15.89 | 15.84 | 20.51% | 22.17% |
| *Government eco. assistance (Q22)* |  |  |  |  |  |  |
| None | 84.59% | 84.13% | 15.33 | 15.34 | 24.76% | 25.32% |
| Any | 15.41% | 15.87% | 14.38 | 14.50 | 30.65% | 31.01% |
| *Activities with perceived low risk of COVID-19 infection (Q21)* |  |  |  |  |  |  |
| Only essential | 10.52% | 10.28% | 13.21 | 13.20 | 41.56% | 41.73% |
| Other activities | 89.48% | 89.72% | 15.60 | 15.63 | 22.63% | 23.13% |
| *COVID-19 Protection Measures (Q26)* |  |  |  |  |  |  |
| None | 2.5% | 3.17% | 18.49 | 18.64 | 16.52% | 17.53% |
| Some | 24.03% | 24.49% | 15.78 | 15.81 | 23.20% | 24.06% |
| All | 73.48% | 72.34% | 14.89 | 14.88 | 26.62% | 27.02% |
| *Psychological Impact (Q31)* |  |  |  |  |  |  |
| None | 30.6% | 34.73% | 15.97 | 16.01 | 22.42% | 23.08% |
| Mild | 13.6% | 13.82% | 15.84 | 15.85 | 20.60% | 20.48% |
| Severe | 55.8% | 51.51% | 14.63 | 14.54 | 28.26% | 29.28% |

Figure 3 shows the results of the average LSNS-6 scores per age group, gender and severity of the economic impact of the pandemic. As seen in the Figure, those who report severe economic impact have significantly lower LSNS-6 scores when compared to the rest (ANOVA, p< 10^−16^, all with posthoc Tukey p< 10^−6^): over 40% of participants with severe economic impact report being socially isolated. This difference in the LSNS-6 scores is particularly significant for female when compared to male respondents of the same age with severe economic impact, as illustrated in Figure 3, bottom. In terms of friends vs family, the LSNS-6 score related to friends is the largest contributor to this very clear gender difference. Regarding age, those aged 60+ with severe economic impact have the lowest LSNS-6 scores of the entire sample: 50.56% of participants in this group report being socially isolated.

**Fig. 3.**
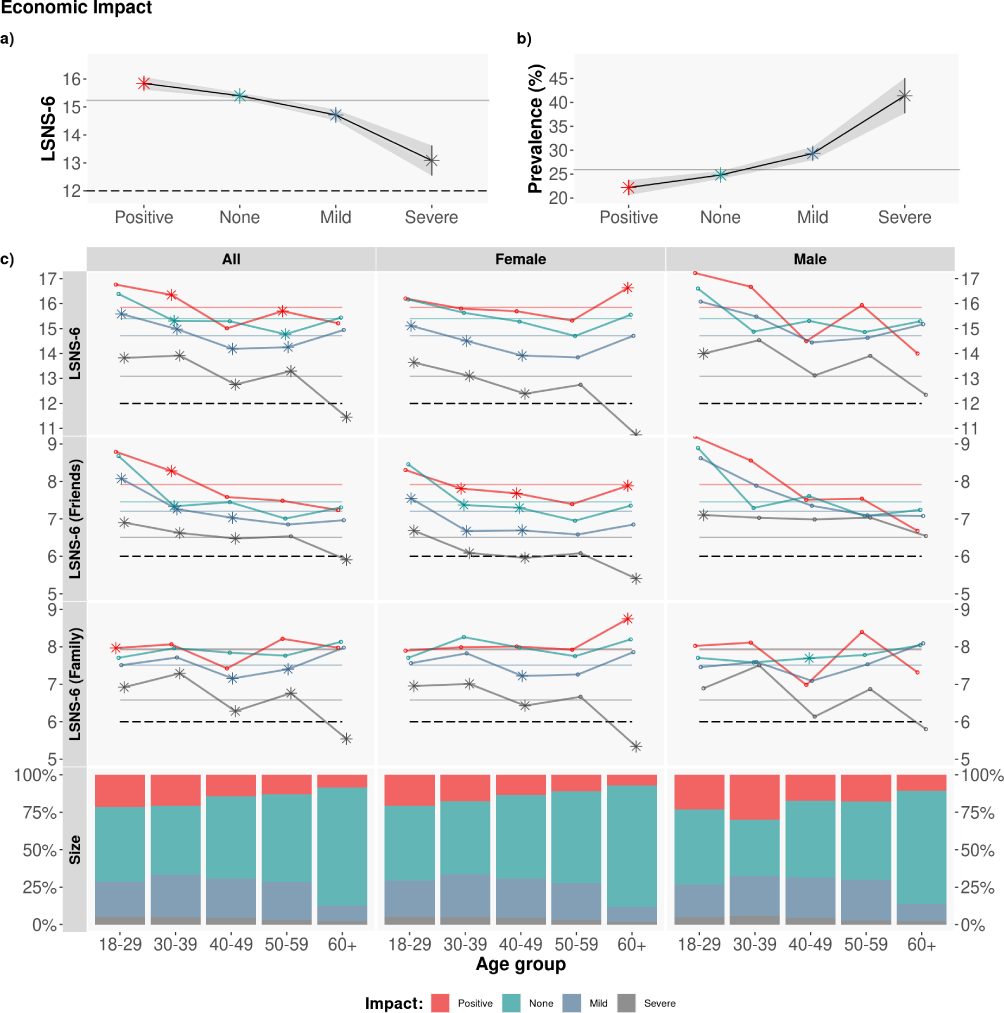
Economic impact. Average LSNS-6 score by age group and gender. The colored dashed lines correspond to the Overall/Friend/Family average LSNS-6 score per economic impact (Positive/None/Mild/Severe): *μ*_*LSNS*−6_ : [15.84, 15.40,14.71, 13.09]; *μ*_*LSNS*−6 *fam*_ : [7.92, 7.94, 7.51, 6.58]; *μ*_*LSNS*−6 *fri*_ : [7.92, 7.45, 7.20, 6.51]. To test the statistical significance of the differences, we applied a Tukey posthoc test after obtaining a significant difference via an ANOVA test. Asterisks denote statistically significant differences with the rest of groups with *α* = 0.05.

Logistic regression analysis confirmed associations economic impact with social isolation in our sample, as illustrated in Table 2.

In addition to the type of economic impact, Q22 in the survey asks whether respondents have received any kind of government economic assistance in the past month. Figure 4 depicts the LSNS-6 scores for those who report receiving government support (red lines) vs those not receiving any type of support by the government (grey lines) per age group and gender. As shown in the Figure, the average LSNS-6 scores of those receiving government support are significantly lower than the scores of those who do not report receiving any kind of support (*μ*_*LSNS*−6_ : 14.50 vs 15.34 for those receiving government support vs not, t-test *p* < 10^−9^ with a difference in means of 0.836).

**Fig. 4.**
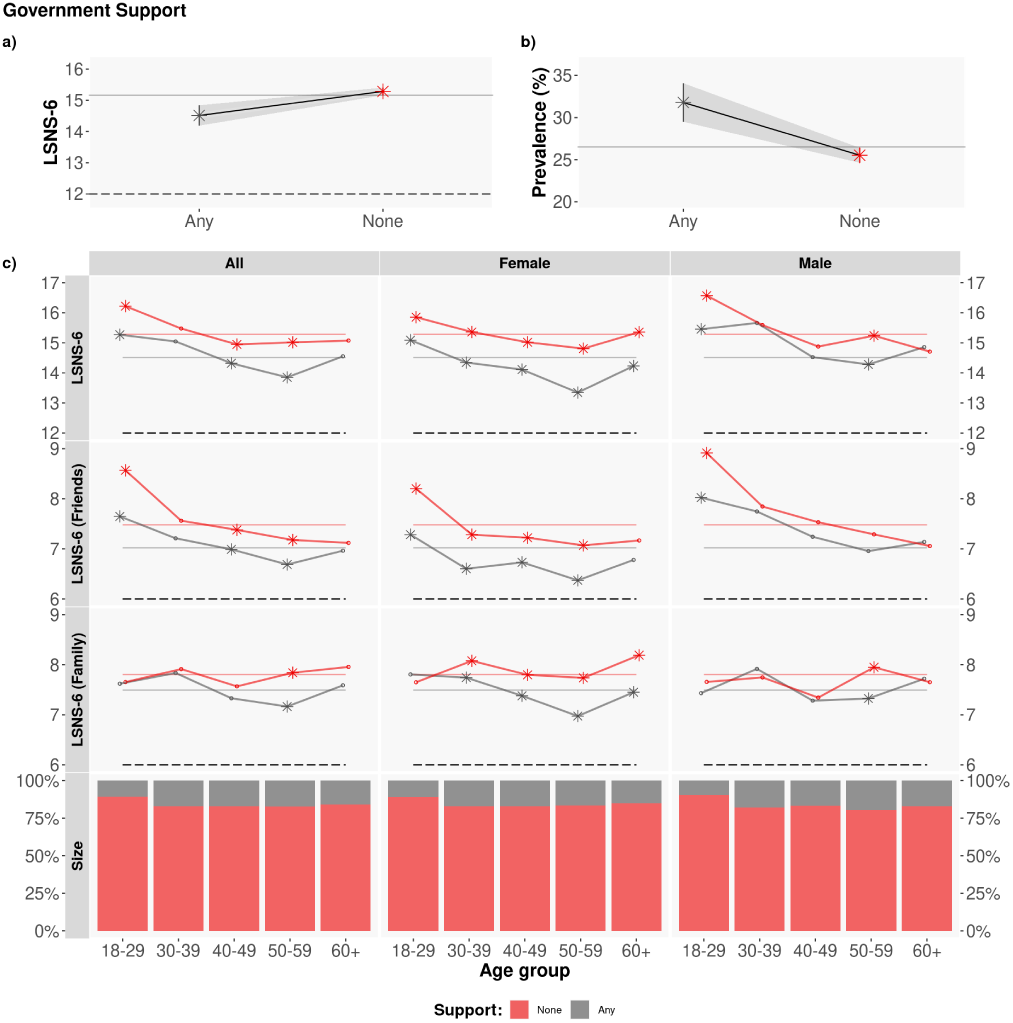
Government support. Average LSNS-6 score by age group and gender. The colored dashed lines correspond to the Overall/Friend/Family average LSNS-6 score per government economic assistance (Any/None): *μ*_*LSNS*−6_: [14.50, 15.34]; *μ*_*LSNS*−6 *fam*_: [7.47,7.77]; *μ*_*LSNS*−6 *fri*_: [7.03, 7.55]. The asterisks denote statistically significant differences, via a t-test with *α* = 0.05.

The largest differences are found in the friend component of the LSNS-6 score, particularly in female respondents. In terms of age, the lowest average LSNS-6 scores correspond to those who receive government support and are 50 to 59 years old, where 35.35% of individuals in this group report being socially isolated.

### RQ3: Social isolation and the psychological impact of the pandemic

We address RQ3 by analyzing the answers to question Q31 in the survey (see Tables 3 and 4 in the S.I.). Figure 5 shows the average LSNS-6 scores per age group, gender and severity of the self-reported psychological impact of the pandemic (*μ*_*LSNS*−6_ : None/Mild/Severe: [16.01, 15.85, 14.54]). Similarly to the economic impact, those who report severe psychological impact have significantly lower LSNS-6 scores than the rest of the respondents. Interestingly, we do not identify significant differences by gender or by type of social support (friends or family) among those who report severe psychological impact. The lowest average LSNS-6 scores correspond to those aged 50 to 59 years old, with 29.54% of members of this group reporting being socially isolated, and 34.90% among those who report severe psychological impact. Logistic regression analysis confirmed associations of psychological impact with social isolation in our sample, as depicted in Table 2, yet milder than the associations found with economic impact.

**Fig. 5.**
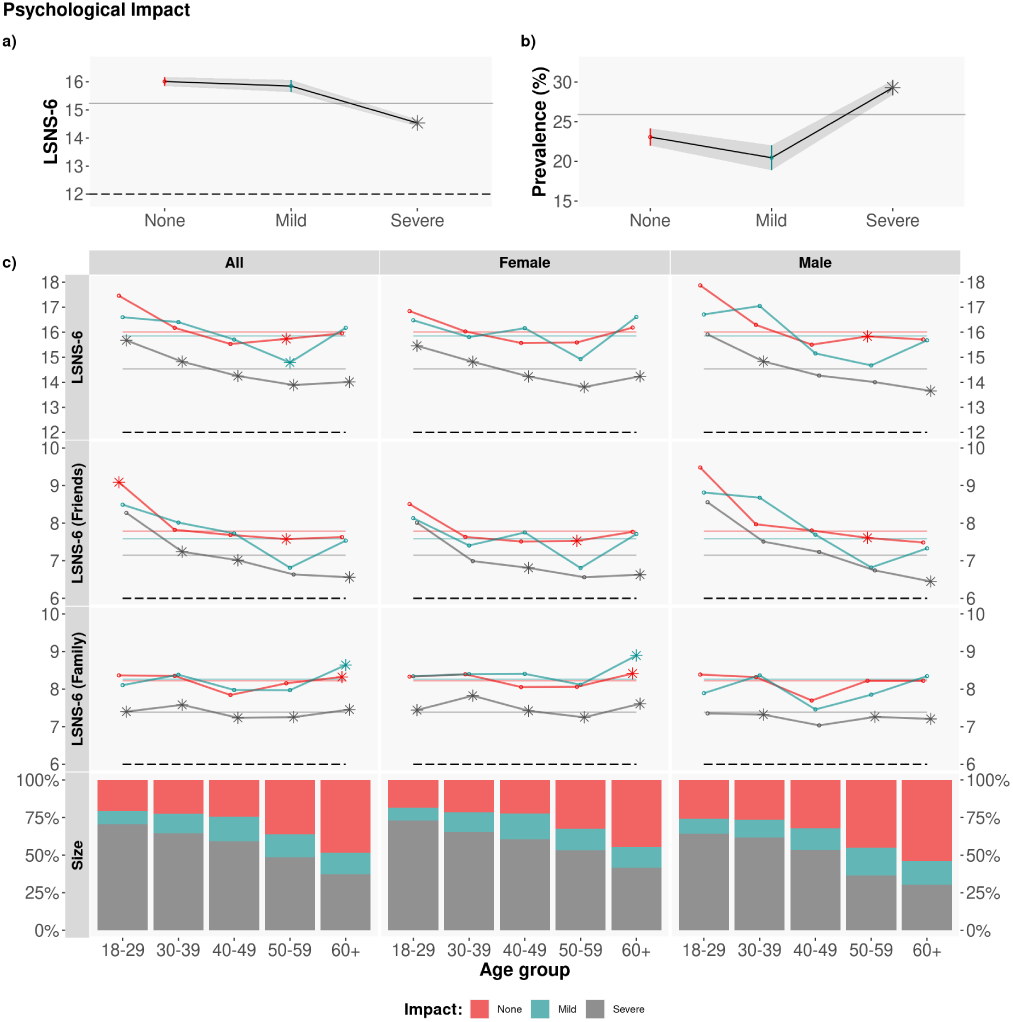
Psychological impact. Average LSNS-6 score by age group. The colored dashed lines correspond to the average overall/friend/family LSNS-6 score depending on the psychological impact (None/Mild/Severe): *μ*_*LSNS*−6_: [16.01, 15.85, 14.54]; *μ*_*LSNS*−6 *fam*_: [8.23, 8.26, 7.39]; *μ*_*LSNS*−6 *fri*_: [7.79, 7.59, 7.15]. To test the statistical significance of the differences, we applied a Tukey post-hoc test after obtaining a significant difference via an ANOVA test. Asterisks denote statistically significant differences with the rest of groups with *α* = 0.05.

### RQ4: Social Isolation, Behaviors and Perceptions related to COVID-19 measures

The fourth research question focuses on the relationship between social isolation and the respondents’ behaviors and perceptions regarding the COVID-19 protection measures that they adopt (Q26) and the perceived risk of coronavirus infection related to different daily-life activities (Q21). The questions and possible answers are described in Tables 3 and 4 in the S.I.

As noted in Figure 6, there is a significant difference in the LSNS-6 scores between those who do not adopt any protection measures (red lines) and the rest of participants (blue and gray lines), with a t-test *p* < 10^−16^ and a difference in means of 3.528. This difference is mainly due to the friends component −− of the LSNS-6 score: those who do not adopt any COVID-19 protection measures report significantly larger LSNS-6 scores than the rest of respondents, i.e. are significantly less likely to be socially isolated: *μ*_*LSNS*− 6 *none*_ −*μ*_*LSNS* −6 *any*_ = 3.528 = 2.408 (friends) + 1.120 (family), with t-test *p* < 10^−9^.

**Fig. 6.**
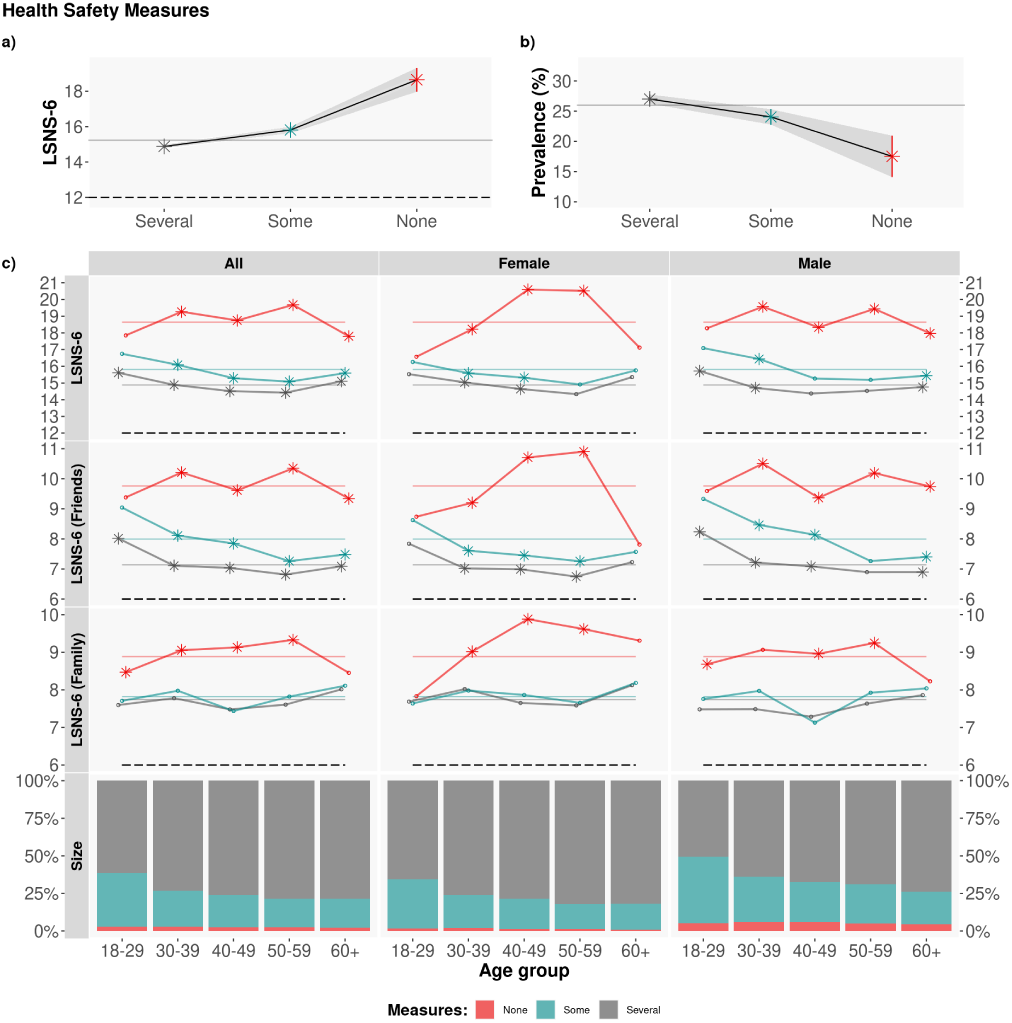
Adopted COVID-19 protection measures. Average LSNS-6 score by age group and gender. The colored lines correspond to the average Overall/Friend/Family LSNS-6 score per adopted COVID-19 protection measures (Several/Some/None): *μ*_*LSNS*−6_: [14.88, 15.81, 18.64]; *μ*_*LSNS*−6 *fam*_: [7.74, 7.82, 8.88]; *μ*_*LSNS*−6 *fri*_: [7.14, 7.99, 9.76]. To test the statistical significance of the differences, we applied a Tukey post-hoc test after obtaining a significant difference via an ANOVA test. Asterisks denote statistically significant differences with the rest of groups with *α* = 0.05.

Next, Q21 asks participants about their perception of COVID-19 infection risk associated with activities that are part of our daily lives. As depicted in Figure 7, there is a significant difference in the LSNS-6 scores between those who report that none or only essential activities can be carried out with low risk of contracting COVID-19 (grey lines) and the rest of respondents (red lines), being this difference mainly due to the friends component of the LSNS-6 score: *μ*_*LSNS*−6 *none*−*essential*_ −*μ*_*LSNS*−6 *other*_ = 2.436 = 1.586 (friends) + 0.850 (family), with t-test p-values < 10^−16^.

**Fig. 7.**
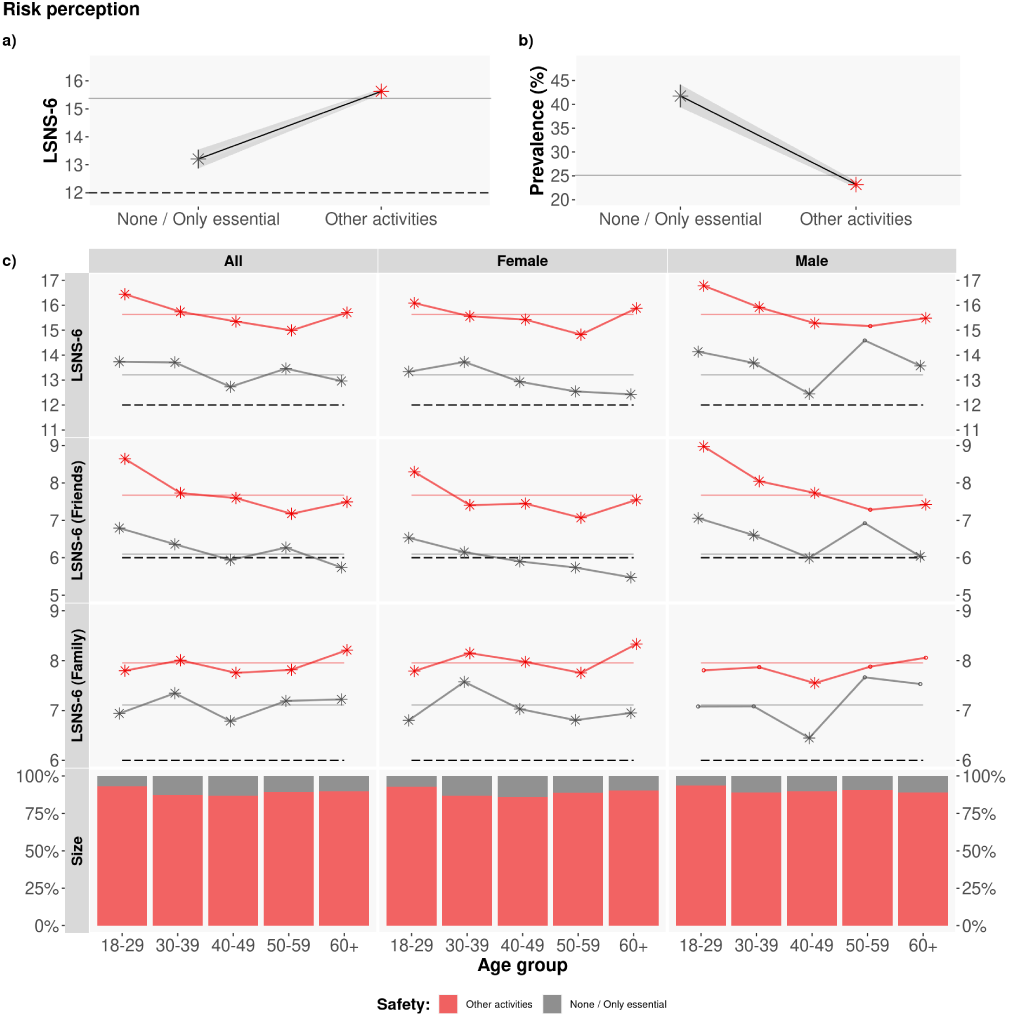
Perception of COVID-19 infection risk for different types of activities. Average LSNS-6 score mean by age group and gender. The colored lines correspond to the average Overall/Friend/Family LSNS-6 score per adopted COVID-19 protection measures (None/Other): *μ*_*LSNS*−6_: [13.20, 15.63]; *μ*_*LSNS*−6 *fam*_: [7.10, 7.95]; *μ*_*LSNS*−6 *fri*_: [6.09, 7.68]. The asterisks denote statistically significant differences, with a t-test with *α* = 0.05.

## Discussion and Implications

In this paper, we have quantitatively analyzed the prevalence of social isolation in Spain during 7 months of the COVID-19 pandemic by means of the LSNS-6 instrument delivered via an online survey with 32,359 answers. We have also studied the relationship between social isolation and the economic and psychological impact of the pandemic on people’s lives. Finally, we have reported the relationship between social isolation, and the COVID-19 protection measures and the perception of infection risk associated with common daily activities by our participants. From our results, we draw several implications.

### The levels of social isolation have significantly increased during the pandemic

While we do not have data of the prevalence of social isolation as per the LSNS-6 instrument in our sample before the pandemic, the European Quality of Life Survey published in 2017 (43) reports that 78% of European adults and 83.5% of Spanish adults have face-to-face contact at least once a week with their family and friends. According to this report, 75% of European adults and 80% of Spanish adults have telephone or internet contact with their social network at least once a week.

Beyond Spain, a recent German study analyzing data from a sample of 9,392 adults (aged 18-79 years old) collected between August 2011 and November 2014 reports a prevalence of social isolation in their sample of 12.3% (30). The average LSNS-6 score in their sample was 17.6 (SD = 5.1). Those socially isolated had a mean LSNS-6 score of 8.5 (SD = 2.5) and the socially integrated individuals of 18.8 (SD = 4.0).

In our sample, we find a prevalence of social isolation of 25.98% (SD = 0.003), which is almost 14 percentage points larger than that reported in (30). The average LSNS-6 score in our sample is of 15.23. Respondents who are socially isolated have an average LSNS-6 score of 7.66 and those socially integrated of 17.89. These figures are lower than those reported in the literature.

Our findings are aligned with those of O^*t*^Sullivan et al.(35), who report a prevalence of social isolation of 21% in a study with 14,302 participants from different countries carried out between June and November of 2020. The authors find that 13% of the respondents in their survey experienced a substantial increase in social isolation during the COVID-19 pandemic.

Moreover, we find a statistically significant difference in the prevalence of social isolation among participants who report not adopting any COVID-19 protection measures –and hence do not reduce their social contacts and activities, (*prev*_*iso*_ = 17.52%) when compared to participants who report complying with five or more COVID-19 protection measures (*prev*_*iso*_ = 27.02%, *χ*^2^ test p< 10^−8^).

This result probably reflects the fact that many of the COVID-19 protection measures are of social nature –such as reducing close contacts, keeping social distance, avoiding gatherings, hugging and kissing– and thus their impact on social isolation is evident.

Hence, we hypothesise that the increase in the social isolation identified in our study is due to the pandemic and the social distancing and confinement measures that have been implemented in Spain to try to contain the spread of the SARS-CoV-2 virus.

### Middle-aged participants are the most socially isolated

Surprisingly, the largest prevalence of social isolation in our sample is among those aged 50-59 years old, with 29.54% of the sample reporting LSNS-6 levels corresponding to social isolation. The LSNS-6 component that contributes the most to this result is that related to the friends network. This finding might be a consequence of the social distancing and confinement measures adopted by Spain during the period of study. According to our survey, the demographic group of those aged 50-59 years old is among the most compliant with the individual protection measures (washing hands, keeping social distancing, wearing masks, limiting social contacts) while also being a demographic group with a large number of contacts. Hence, the impact of adopting such measures on their friends social network might have been the largest in the sample.

Previous work has obtained similar results in smaller samples and geographies. In a small sample of 214 residents of Wandsworth, a South West London Borough in the United Kingdom (34), the authors find that middle-aged people reported a less strong social network and more loneliness, anxiety and depression than younger people. Sugaya et al. carry out a study with 11,333 participants from seven prefectures of Japan during the final phase of the state of emergency in May 2020 and report a greater prevalence of social isolation among male, middle-aged (4064 years) participants (32).

Interestingly, the U-shaped curve in the LSNS-6 scores that we obtain is similar to the previously reported U-shaped curve of happiness depending on age (44) where the lowest levels of happiness are reported by those aged 50 years old.

Given the strong correlation between social isolation and mental and physical health, it would be of paramount importance to deploy support programs for individuals, particularly in such an age group.

### Economic impact and social isolation are strongly related

Previous work has also found a strong relationship between socio-economic status and social isolation both before (5, 8, 40, 45–48) and during the COVID-19 pandemic (30, 32, 34, 49–51). The general finding that we corroborate here is that the lower the socio-economic status, the higher the risk for social isolation.

The economic consequences of the COVID-19 pandemic are evident on a global scale, having been responsible for the second largest global recession in history, with more than a third of the global population at the time being placed on lockdown.

However, we are not aware of previous analyses on the relationship between the economic *impact* of the pandemic on an individual and their levels of social isolation. In our data, we find a strong correlation between the economic impact of the pandemic and the LSNS-6 scores of individuals: the lower the economic impact, the larger the LSNS-6 scores (see Table 2). The prevalence of social isolation among those who report severe economic impact is significantly larger than the prevalence of the overall sample: *prev*_*iso*_ = 41.39% and *prev*_*iso*_= 25.98% for those with severe economic impact vs the general population, respectively.

We also observe gender differences, mainly due to the friends component of the LSNS-6 scores: women who have been severely impacted economically because of the pandemic are more likely to be socially isolated (*prev*_*iso*_ = 43.04%) than men who also report severe economic impact (*prev*_*iso*_ = 39.78%), with average friend LSNS-6 scores almost one point smaller than the men (*μ*_*LSNS*−6 *fri*−*female*_: 6.04 *μ*_*LSNS*−6 *fri*−*male*_:6.96).

This finding highlights the importance of deploying social programs to support women in vulnerable and economically precarious situations.

### The social support by friends matters

We find that the friends component of the LSNS-6 score is a key determinant of social isolation in our sample across all age groups (see Figure 1 d), more so than the family component. This finding might be due to the fact that families are a fundamental pillar in Spanish culture across all demographic and socio-economic groups, such that there might not be large differences in the social support provided by families. However, the friends networks vary widely with age, gender, education level and socio-economic status (52).

We identify gender differences in the levels of social isolation for those who report severe economic impact due to a much weaker friends social network among women.

There are also significant differences in the friends LSNS-6 scores among those who do not adopt any COVID-19 protection measures (particularly men) and the rest of participants; and those who consider that most activities can be performed with low risk of a SARS-CoV-2 infection and the rest of respondents. In this case, those who do not adopt any COVID-19 protection measures exhibit stronger social support by their friends. Similarly, those who consider that many non-essential activities can be carried out with a low risk of coronavirus infection also have higher LSNS-6 friend scores than the rest of participants.

This finding highlights not only the role that friends play in social isolation – particularly among the youth (see e.g. (53)), but also the influence that having a strong network of friends plays on wanting to socialise even during pandemic times. Alternatively, this result might be explained by the fact that the COVID-19 protection measures are mostly of social nature and hence have an impact on the levels of social isolation of those who comply with them.

### Gender also matters

There are several gender-based differences that can be drawn from our analyses. First, while the average LSNS-6 scores are lower among female (*μ*_*LSNS*−6_ = 15.18 *SD* = 0.048) than male (*μ*_*LSNS*−6_ = 15.29 *SD* = 0.074) respondents, the prevalence of social isolation is larger among male (*prev*_*iso*_ = 26.53%) than female (*prev*_*iso*_ = 25.47%) participants. This difference, however, is not statistically significant according to a *χ*^2^ test.

To shed light on this finding, Figure 9 in the S.I. depicts the distribution of LSNS-6 scores by gender. As can be observed in the Figure, the LSNS-6 scores by female respondents tend to be clustered around the mean of the distribution whereas the upper and lower extremes of the distribution have significantly more male than female respondents.

Second, women with severe economic impact are significantly at higher risk of social isolation than men who have also suffered severe economic impact, mostly due to having a weaker support by their friends. Interestingly, this difference is not evident among those with psychological impact due to the pandemic.

### The adoption of COVID-19 protection measures is related with social isolation

When we divide our sample according to the COVID-19 protection measures adopted by the participants in our study, significant differences emerge in the respondents’ levels of social isolation: those who report not adopting any measures have significantly larger LSNS-6 scores than those who adopt some or all of the possible COVID-19 protection measures (None/Some/Several: *μ*_*LSNS*−6_: [18.65, 15.81, 14.88], ANOVA and Tukey tests *p* < 10^−16^.

This finding might be explained by the role that the friends network plays in influencing human behavior. Many of the COVID-19 protection measures are meant to limit our social interactions. Those with strong friends networks are therefore more impacted by the adoption of such measures and hence could be less likely to adopt them. As previously reported, the difference in the LSNS-6 scores between those who do not adopt any measures and the rest of participants is of 3.524 points, mostly due to the friends component of the LSNS-6 score.

Alternatively, this result might be explained by the fact that the COVID-19 protection measures are mostly of social nature and hence have an impact on the levels of social isolation of those who comply with such measures.

### Higher perception of COVID-19 infection risk is related to social isolation

Similarly, those who report that many daily activities can be performed with low risk of a coronavirus infection are significantly less likely to be socially isolated than those who have a higher perception of infection risk: *prev*_*iso*_ = 23.13% vs *prev*_*iso*_ = 41.73% (*χ*^2^ test with *p* < 10^−16^). Again, the LSNS-6 component that contributes the most to this difference in LSNS-6 values (13.20 vs 15.63) is the friends component: from the 2.44 points in the difference of the average LSNS-6 scores, 1.59 points are due to the friends component and 0.85 points due to the family component, with a t-test *p* < 10^−16^, and with a larger difference between female than male respondents. Our results might be explained by several factors. First, previous work has found that the larger the social support, the lower the fear towards COVID-19 in a sample of healthcare workers in Jordan (54). Second, in the context of the coronavirus pandemic, we tend to lower our perception of infection risk when carrying out activities that involve our friends and/or relatives(55): “our evaluation of risk versus safety is inextricably tied to our group memberships”.

Several of our findings highlight the crucial role that the friends support network plays in the levels of social isolation in Spain.

## Materials and Methods

### Data Collection and Processing

In this paper, we analyze a subset of the answers to the COVID19ImpactSurvey, an extensive, anonymous, online citizen survey about COVID-19 (36). Launched on March, 28th 2020, in Spain, the survey has since then collected over 700,000 anonymous answers from 11 countries (mostly Spain, Italy, Germany and Brazil), and it is available online at https://covid19impactsurvey.org. Participants must declare being 18 years or older to be able to fill the survey. All research was performed in accordance with relevant guidelines/regulations. Informed consent was obtained from all users and the data was collected in de-identified form.

The specific answers to the survey that we analyze in this paper collect information about the participants’ demographic and household situation (Q1-Q6); their LSNS-6 scores (Q11-Q16); the economic impact of the pandemic in their lives (Q20, Q22); their perception of COVID-19 infection risk associated with different activities (Q21); the individual protection measures that they adopt to protect themselves against COVID-19 infections (Q26); the psychological impact of the pandemic in their households (Q31). All the questions to the survey are included in Table 3 in the S.I.

We analyze the data from Spain for the time period between June 6th, 2021 and the 16th of December, 2021. During this period, Spain had returned to the “new normality”: the only COVID-19 protection measure implemented in the country was the requirement to wear face masks in indoor public spaces. Survey answers were collected from volunteer respondents who learned about the survey via social media channels, word-of-mouth, universities and news organizations. We used Facebook and Instagram ads as an additional channel to recruit volunteers. This approach gave us a straightforward method to obtain a broad sample of users across different socioeconomic and demographic groups. We did not use any targeting feature except for gender, where we used separate budgets to balance the numbers of male and female respondents. The cost-per-successful-response was €0.24 and €0.11 for men and women respectively. Both men and women had similar completion rates (54%) once they began answering the survey.

The age and gender distribution of our sample might be different from the population distribution in Spain. Thus, we follow the methodology described in Oliver *et al*. (36) and re-weight the answers such that the resulting gender and age distributions match the official statistics of each of the seven Nielsen geographical regions in Spain, depicted in Figure 8, plus the metropolitan areas of Madrid and Barcelona. We compute the weights according to the official data reported by the Spanish National Institute of Statistics (INE) for 2020.

**Fig. 8.**
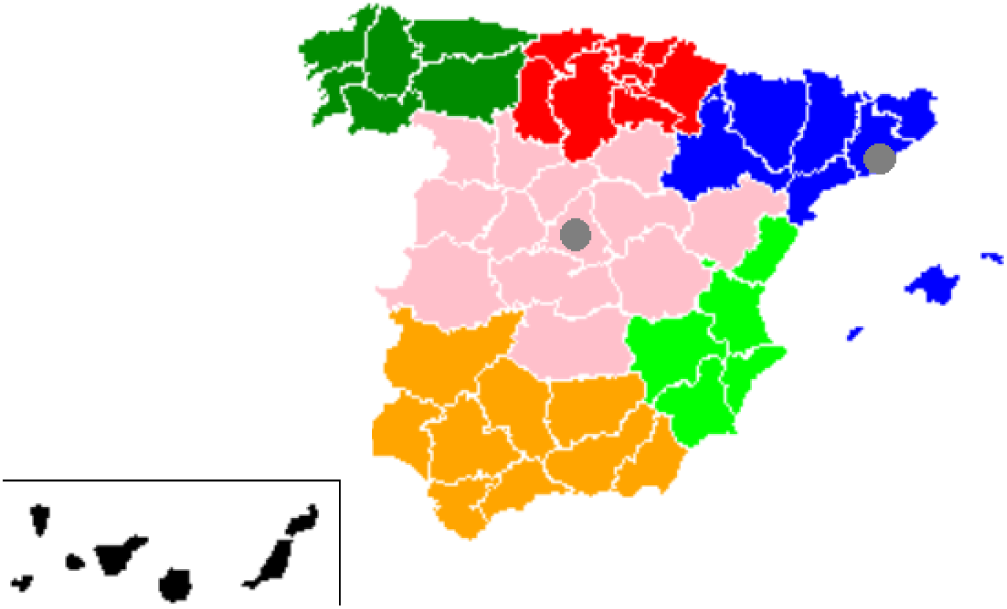
The 7 Nielsen areas plus the metropolitan areas of Madrid and Barcelona, used to reweigh the data.

**Fig. 9.**
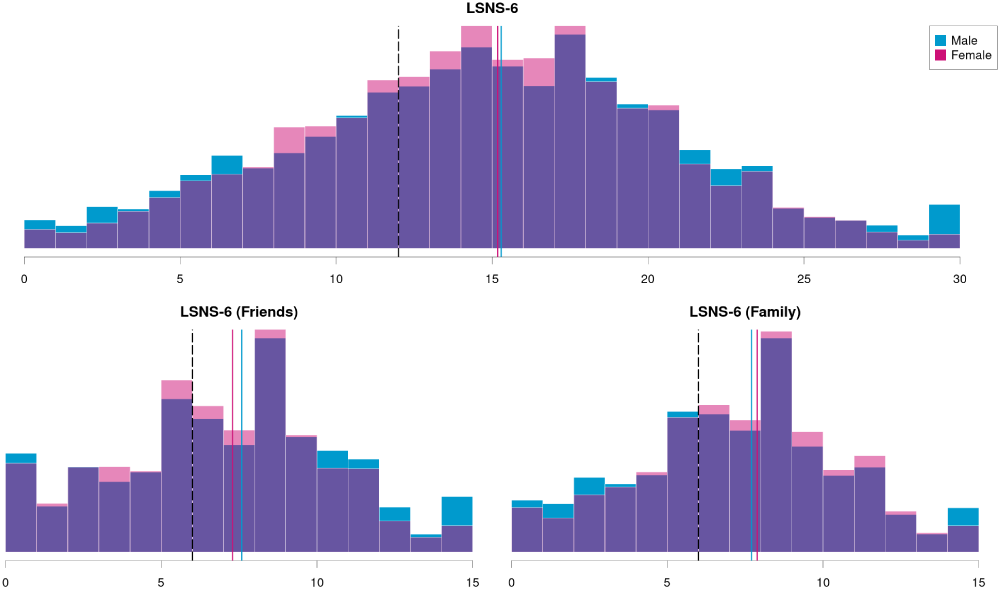
LSNS-6 scores distribution by gender: blue male pink female, the vertical lines show the mean LSNS-6 by gender, and the dash vertical lines shows the isolation level LSNS-6 <12

To further validate our methodology, in April 2020 we commissioned an IPSOS.digital FastFacts panel of a cohort of 1,000 representative general population respondents aged 18-65 in Spain. This validation was done after we started using Facebook ads, but before the time period covered by this paper. The results of the IPSOS panel were within the margin of error of our survey results for the same time period and ages.

#### Ethical Approval

All methods described in this manuscript were carried out in accordance with relevant guidelines and regulations. The survey instrument was approved by the Ethical Committee of the University Miguel Hernandez, which is called the Office for Responsible Research (OIR), in the context of previous research (36, 37). Informed consent was obtained from all subjects who had to confirm being at least 18 years old to be able to participate in the survey. The participation in the survey was voluntary and all the collected data was anonymous and confidential. Furthermore, the research complied with the ethical standards of the 1964 Declaration of Helsinki and its later amendments.

### Measures

All the measures used in our analysis are described in Tables 3 and 4 of the S.I.

#### Social Isolation

We measure social isolation using the LSNS-6 instrument included in questions Q11 to Q16 in the survey. The LSNS-6 is a quantitative measure of an individual’s social network size that assesses both the number and frequency of contacts with friends and family and the social support received by them. It consists of six items: three questions ask about the frequency of meeting relatives, how many relatives the respondent feels close enough to ask for help and with how many relatives the respondent can talk about private matters; the other three questions are identical but asking about friends rather than relatives. Each of the LSNS-6 items is scored from 0 to 5 and they are all equally weighted. Thus, the total score ranges between 0 and 30. The higher the LSNS-6 scores, the lower the levels of social isolation. Social isolation is captured by a score below 12, as it means that on average there are fewer than 2 individuals available to the respondent in the areas probed by the survey (i.e. family or friends). The LSNS-6 has good psychometric properties (13).

#### Socio-demographic and household characteristics

Demographic variables selected for analysis were: (1) gender (Q2 in the survey), respondents were asked to self-report their gender as male, female or other. In this study, we only consider the answers of those who responded as male or female, which represent 99.33% of the answers (0.3% of answers selected “other” as their sex and 0.37% of answers did not provide sex information); (2) age (Q1 in the survey), coded in the following bins: 18-29; 30-39; 40-49; 50-59; 60+; (3) zip code of their residence (Q3 in the survey); (4) number of members in the household (Q5 in the survey), coded as: 1, 2, 3, 4+.

### Economic impact

The self-reported economic impact of the pandemic was assessed in Q20 and Q22 of the survey.

Question number 20 asks about the type of economic impact that the pandemic had on the respondents’ lives. Based on the answers provided to Q20, we coded the severity of the economic impact in four levels: [None, Mild, Severe and Positive], as described in Table 4 in the S.I.

Question number 22 focuses on whether participants receive any kind of economic assistance from the government. Based on the answers provided to Q22, we coded the reliance on government-based economic assistance in two levels: [None, Any], as described in Table 4 in the S.I.

#### Psychological impact

The self-reported psychological impact of the pandemic was assessed in Q31 of the survey. Based on the answers provided to Q31, we code the severity of the psychological impact in three levels: [None, Mild and Severe], as described in Table 4 in the S.I.

#### COVID-19 protection measures

The individual protection measures adopted by participants to limit their exposure to the SARS-CoV-2 virus were captured by Q26. We code the possible answers into three groups: [All, Some, None], depending on whether respondents report adopting all, none or several of the measures, as described in Table 4 in the S.I.

#### Perception of COVID-19 infection risk

The participants’ perception of risk of a COVID-19 infection associated with different daily-life activities was the focus of Q21 in the survey. We code the possible answers into three groups: [None, Only essential activities and Other non-essential activities], as depicted in Table 4 in the S.I.

### Statistical analysis

The prevalence of social isolation is computed in percent with 95% confidence intervals (CIs) from the overall sample. We stratify social isolation by gender and age. We perform *χ*^2^ tests to determine the existence of statistically significant differences in the prevalence of social isolation among each of the stratified groups. We also use multi-variable logistic regression analysis to test the associations between social isolation and age, gender, number of members of the household, economic and psychological impact of the pandemic, individual COVID-19 protection measures adopted and risk perceptions of coronavirus infection associated with different daily activities. We use a significance level *α* = 0.05 (two-tailed) in all the analyses.

We carried all our statistical work using the R software package.

### Limitations

Collecting a large sample of answers via an online survey does not come without limitations. Even if the data size is large and the confidence intervals are small, errors might compound and under- or over-estimate the results (56). Moreover, even if we weighted (57) the data to mitigate the survey’s gender and age biases in each Nielsen region, and we deployed gender-balanced Facebook advertisement campaigns, the data is non-random and biases might still be present. For example, there are self-selection and sampling biases (58) as the survey is filled out by volunteers who have learned about the survey via social media, WhatsApp, newspapers’ articles or Facebook ads, and who need to have access to a computing device (smartphone, tablet, PC) with an Internet connection. The survey might also be biased towards people who are more likely to fill-in surveys, such as young and highly educated people. Respondents must be adults –at least 18 years old. Hence, students are only partially covered. We acknowledge all these biases challenge the estimation of inferential statistics, and might lead to overconfidence in incorrect inferences due to the collected large data sample and small confidence intervals (56). We also acknowledge that there might be a recall bias as respondents were asked about their perceptions and behavior in the last seven days (e.g. the number of close contacts in the last week).

However, we have taken several measures (re-weighting, contrasting with two polls, comparing with relevant literature) to minimize such biases. Previous work has found that citizen (online) surveys are particularly valuable tools in situations of data scarcity where informed and timely decisions are needed (36). Online surveys also allow monitoring people’s perceptions and behaviors, enabling the design of more effective public policies and better education and communication with the public. Thus, we are confident about the value and validity of the results of our analyses.

## Data Availability

All data produced in the present study are available upon reasonable request to the authors

https://ellisalicante.org/en/covid19impactsurvey

## Author contributions

N.O. and K.R. designed, deployed and conceived the study; N.O., M.MG. and E.S. defined the methods; M.MG. and E.S. conducted the statistical analyses; R.F. helped with the official statistics data; N.O., M.MG. and E.S. analyzed the results; A.CH. performed the literature review; N.O. wrote the manuscript, which was reviewed by all authors.

## ACKNOWLEDGMENTS

N.O. has been partially supported by funding received by the ELLIS Unit Alicante Foundation from the Regional Government of Valencia in Spain (Generalitat Valenciana, Conselleria d’Innovació, Universitats, Ciència i Societat Digital, Dirección General para el Avance de la Sociedad Digital), by virtue of a collaboration agreement signed in 2021 (Convenio Singular 2021). N.O., M.MG., E.SS. and R.F. have been partially funded by a grant from the BBVA Foundation through the IA4COVID19 research project.

## Supporting Information

### Survey questions

Table 3 depicts the questions whose answers have been analysed in this paper. The survey has a total of 31 questions. However, there are a few conditional questions such that the number of the questions is that a person might answer could be more than 31. There are gaps in the numbering since during the lifetime of the survey questions regarding the lockdown behaviour were removed as they were no longer relevant.

Table 4 depicts the dependent and independent variables used in this study.

